# Deciphering early-warning signals of SARS-CoV-2 elimination and resurgence from limited data at multiple scales

**DOI:** 10.1101/2020.11.23.20236968

**Authors:** Kris V Parag, Benjamin J Cowling, Christl A Donnelly

## Abstract

Inferring the transmission potential of an infectious disease during low-incidence periods following epidemic waves is crucial for preparedness. In such periods, scarce data may hinder existing inference methods, blurring early-warning signals essential for discriminating between the likelihoods of resurgence versus elimination. Advanced insight into whether elevating caseloads (requiring swift community-wide interventions) or local elimination (allowing controls to be relaxed or refocussed on case-importation) might occur, can separate decisive from ineffective policy. By generalising and fusing recent approaches, we propose a novel early-warning framework that maximises the information extracted from low-incidence data to robustly infer the chances of sustained local-transmission or elimination in real time, at any scale of investigation (assuming sufficiently good surveillance). Applying this framework, we decipher hidden disease-transmission signals in prolonged low-incidence COVID-19 data from New Zealand, Hong Kong and Victoria, Australia. We uncover how timely interventions associate with averting resurgent waves, support official elimination declarations and evidence the effectiveness of the rapid, adaptive COVID-19 responses employed in these regions.

## Introduction

The timeliness of the application and relaxation of non-pharmaceutical interventions (NPIs) (e.g., border closures, quarantines or social-distancing mandates) has been a polarising and pressing topic of global debate throughout the COVID-19 pandemic. Deciding on how best to balance the risk of resurging infections (second or later waves) against the costs (economic and otherwise) of sustaining NPIs and related restrictions is non-trivial and still lacks clear consensus. Among the most widely used early-warning analytics informing NPI policy is the effective reproduction number (*R*) [1,2], popularly displayed on numerous COVID-19-related websites and dashboards [3–6]. While, in theory, an escalation from *R* < 1 (the epidemic is waning) to *R* > 1 (it is growing) forewarns of resurgence, robustly and reliably identifying this transition when case-incidence is small is fundamentally difficult, in practice [7–9].

Low-incidence periods contain necessarily scarce data, which can often hinder standard *R*-inference approaches, limiting their reliability or forcing them to rely excessively on prior assumptions [1,7,10,11]. However, trustworthy disease-transmission estimates during those periods, which are characteristic of the lull between potential epidemic waves for example, are crucial for informing decision-making, providing early indicators for discriminating between the starkly different possibilities of elimination (i.e., no future local cases [2,12]) and resurgence. Inferring transmission dynamics at low incidence has been highlighted as a key challenge to designing safe protocols for NPI relaxation across the pandemic [11].

These problems are only exacerbated by the important and distinct roles of local and imported cases, both in controlling the chances of elimination or resurgence, and in defining effective NPI policy [2]. At low incidence, it is essential to distinguish between (i) true second waves of community transmission, which may necessitate broad-spectrum NPIs e.g., local lockdowns, and (ii) multiple, stuttering epidemic chains seeded by repeated importations, which require targeted NPIs e.g., isolation of travellers. Failing to properly account for local-import dynamics can inflate *R*-estimates, confounding (i) with (ii) and potentially misleading policymakers [13].

Moreover, estimating the likelihood of elimination and hence the endpoint of a local epidemic is non-trivial. While the World Health Organisation (WHO) recommends waiting fixed, disease-specific times (e.g., 28 days for COVID-19), from the last observed case, before declaring an outbreak over [12], this approach is insensitive to variations among incidence curves of the same disease [14] and neglects local-import case distinctions [15]. Recent methods, which better incorporate epidemic data to derive tailored and contextualised measures of elimination, however, are still intrinsically hindered by the poor reliability of *R*-estimates at the epidemic tail [15]. Consequently, more robust, data-driven outbreak analytics are needed to bolster the evidence base for NPI policy and decision-making during critical low-incidence periods [11].

Here we present a novel early-warning framework for robustly assessing *R* and the likelihood of elimination, which circumvents the above problems, highlights the diverse roles of imported and local cases and underscores how well-timed, adaptive NPI application and relaxation can avert resurgence and promote local elimination. Our framework introduces two analytics: the smoothed *local R* and *Z* numbers, which measure community transmission and the confidence in local elimination, respectively, at any time and scale of interest. Our *R* improves on widely-used approaches such as *EpiEstim* [7] and the *Wallinga-Teunis* method [10] by generalising new methodology [16] that solves what is termed the *smoothing* problem in engineering [17], to include the local-import model previously used to investigate (i)–(ii) for zoonoses [18].

*Smoothing* solutions formally maximise the signal extracted from noisy datasets [17,19]. Our *R*-estimates exploit both forward- *and* backward-looking information from a given incidence curve (see Methods). Standard approaches use only forward- [10] *or* backward-looking [7] information, which limits their ability to decipher crucial trends hidden in the data. As a result, our *R*-estimates (Eq.1) can be significantly more robust in low-incidence periods (see Results) and accordingly better at providing reliable, advanced warnings of resurgence. Our *Z* number extends recent methods for forecasting epidemic lifetimes [15] to exploit these smoothed local *R*-numbers and to improve the quantification of uncertainty in their estimates (Eq.2). The result is a meaningful measure of our confidence at any time-point that there will be no future local cases i.e., that the epidemic is eliminated or will fade out.

Our *R-Z* framework can therefore expose transmission signals buried in scarce data to provide early risk-assessments of resurgence or confirmations of elimination. As it only requires local and imported case classifications, this framework can be applied at any scale (e.g., country-wide or sub-regionally) in real-time or retrospectively. We showcase its power by evaluating the alignment of NPI policy and key COVID-19 transmission dynamics in New Zealand, Hong Kong and Victoria state, Australia. While the demographics, epidemic curves and policies in these case-studies differ, all feature prolonged durations of low-incidence and appreciable case-importations that have stymied previous attempts at extracting insight into the interplay among NPIs and transmission potential [1,3–6]. Our analysis strengthens the evidence base for the effectiveness and timeliness of the strategies each location employed.

## Results

We examine three case studies involving local COVID-19 dynamics for New Zealand, Hong Kong and Victoria state, Australia. Our main results are in Figures 1-3. While the *Z* metrics are always computed sequentially in real time, the *R*-estimates shown below are retrospective as we process the entire incidence curve over our study period. This means that they present the most informative view of transmission possible (see Methods). We provide corresponding real time *R*-estimates in the Supplement (Figures A, D and G), which only process portions of the incidence curve up to key intervention timepoints. These analyses largely correspond with Figures 1-3 (usually agreeing within 3 days of additional data), and underscore the benefit of our framework for deciphering key early-warning signals of transmission dynamics.

**Figure 1:**
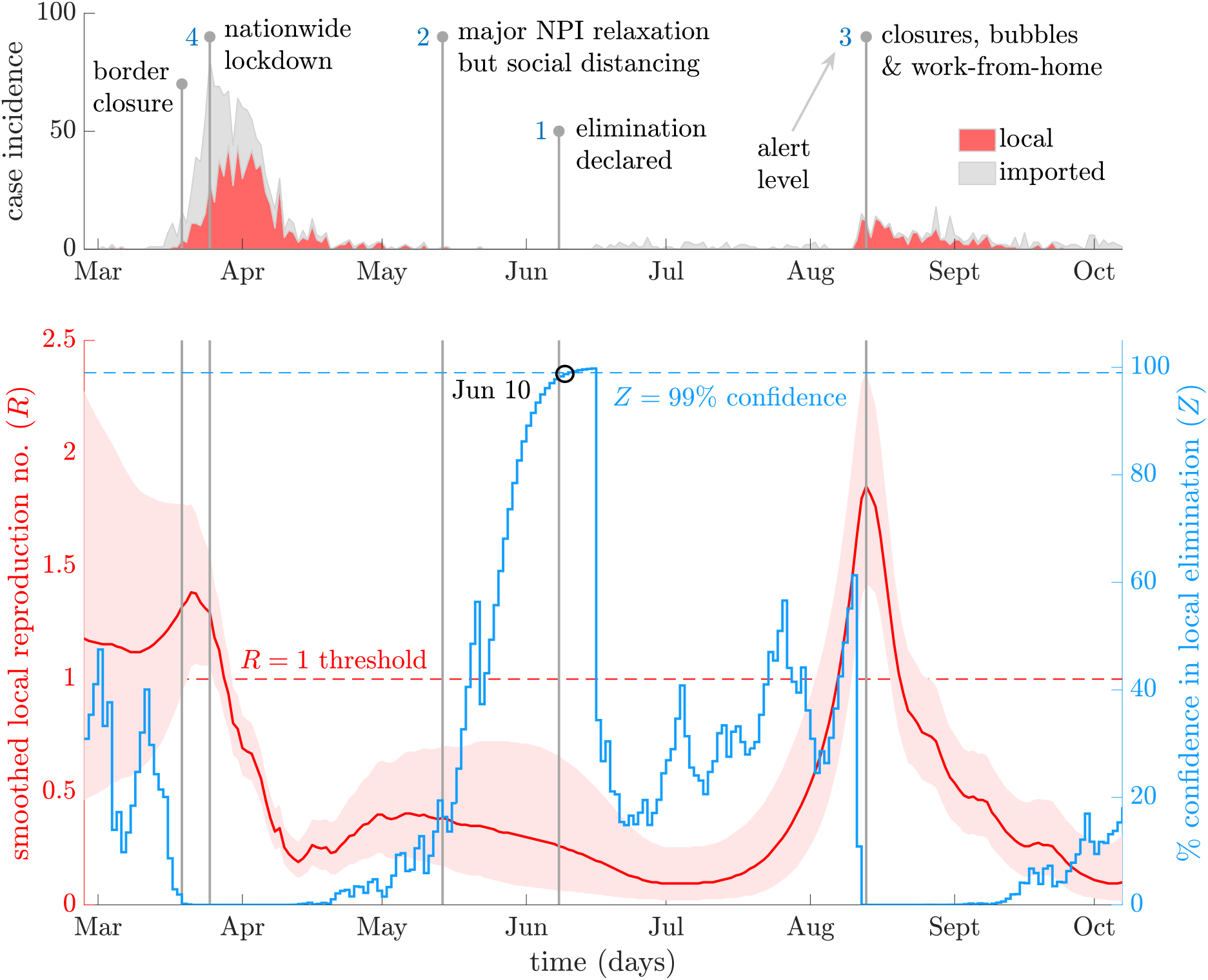
Local transmission dynamics of COVID-19 in New Zealand. The top panel plots local (red) and imported (grey, stacked) cases by date reported, sourced from [21]. Vertical lines pinpoint key policy change-times and alert levels (blue numbers) in response to these caseloads. The bottom panel presents smoothed local *R* number estimates (red with 95% confidence bands) and *Z* numbers (blue), which measure % elimination potential.

### Elimination and import-driven resurgence in New Zealand

New Zealand recorded local transmission of the SARS-CoV-2 virus in mid-March of 2020 and, within 2 weeks, initiated border closures (19 March) and devised a 4-level alert system for NPI deployment, with the aim of elimination [20]. Elevated caseloads quickly culminated in national lockdown (level 4) on March 26, which involved stay-at-home orders and wide venue closure. As the epidemic waned, NPIs were relaxed by May 14 (level 2), although social distancing remained enforced. Subsequently, no cases were observed for a prolonged period leading to a declaration of elimination on June 9 (level 1) [20]. However, local cases were detected again in early August and NPIs (e.g., contact bubbles and work at home orders) were swiftly enacted by August 12 to avoid resurgence (levels 2-3). De-escalation (level 1) followed on October 7; the last date we analyse. Figure 1 (top) summarises this case timeline with data from [21].

Applying our *R*-*Z* approach, we demonstrate how NPI decision-times align with community transmission in Figure 1 (bottom). Initially, there was notable uncertainty around *R* suggesting either supercritical or subcritical transmission could occur. The early response of New Zealand likely suppressed the first possibility, confidently forcing *R* under 1, post-lockdown. Swift action here was potentially critical as delayed responses in other countries have been correlated with larger epidemic sizes. An *R* < 1 was sustained for a significant period after most NPIs were relaxed. Naïve *R*-estimates, which ignore local-import case divisions, would falsely predict *R* > 1 across much of this period (see Supplement). This naïve *R*, which is often presented in COVID-19 analyses and dashboards [1,3], could misinform policymakers.

Post-relaxation, the *Z* number, which characterises risks to elimination from both imported and local cases, increased, suggesting the first wave could be declared over with 99% certainty by June 10. This corroborates the official declaration on June 9 [20]. Subsequently, recurrent introductions seeded new outbreaks, which led to the *R*-estimate climbing confidently above 1 just before the resurgence action-point. This steep rise in *R* (and fall of *Z*) highlights that not only was a second wave likely but also that its transmission potential was larger than the first. The timely, unequivocal response of New Zealand in August likely averted a more explosive second epidemic, correlating with suppressed community transmission. The observed sharp decline in *R* and its remaining below 1 evidences the efficacy of this policy and supports the belief that New Zealand regained control of COVID-19 in early October 2020.

### Partial elimination and multiple waves in Hong Kong, China

Upon learning of the SARS-CoV-2 outbreak in Wuhan, Hong Kong acted quickly, mobilising intensive surveillance schemes and declaring a state of emergency on January 25, 2020 in response to initial cases [22]. This involved closing amusement parks and suspending school reopening, which together with further NPIs enacted throughout February, likely suppressed wave 1. However, wave 2 began in March with many imported cases from North America and Europe, prompting strict border-closures on March 25 and bans on major public gatherings on March 29. Following these and other measures (e.g., venue closures) across April, incidence reduced. Consequently, NPIs were relaxed gradually from May 5-27. While imported cases continued to be recorded 21 days passed with no local cases observed, ending on July 5 [23].

Wave 3 soon surfaced with multiple, local infection-clusters in early July sharply increasing incidence. Consequently, mask mandates and social distancing controls were introduced on July 13 with additional measures enforced by July 19. Further tightening of these measures later in July eventually mitigated the wave, allowing NPI relaxations in September. Incidence was sustained at a low-level for two months before another resurgence occurred as wave 4 on 24 November (with NPIs re-applied), the last date we analyse. Figure 2 (top) plots this timeline, with data from [6]. Although Hong Kong’s response is less discretised than New Zealand’s, our *R-Z* framework still reveals sharp correlations between NPIs and salient transmission dynamics, as illustrated in Figure 2 (bottom).

**Figure 2:**
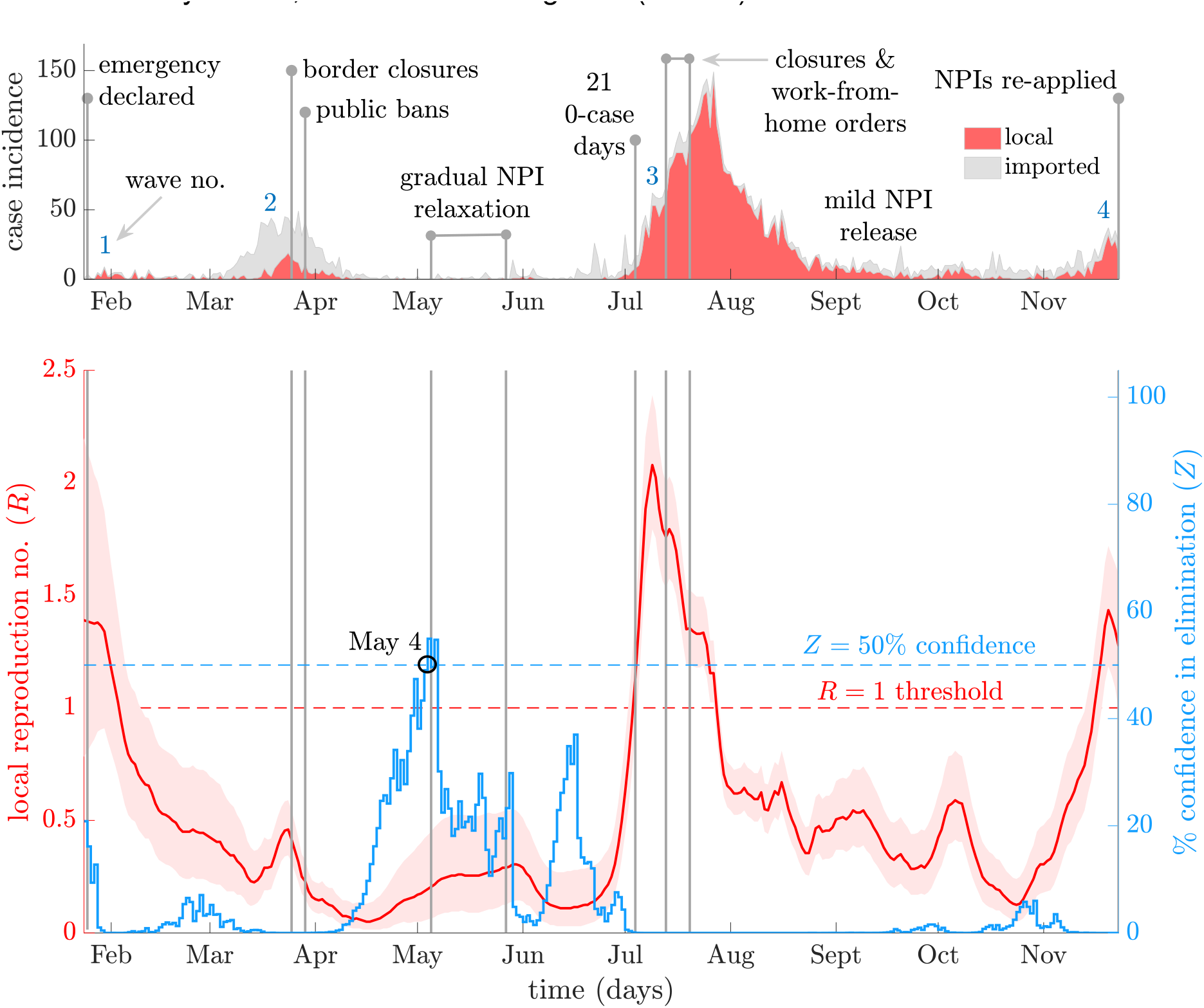
Local transmission dynamics of COVID-19 in Hong Kong, China. The top panel presents local cases (red) and imported cases (grey, stacked) by onset date from [6]. Vertical lines demarcate key policy change-times and responses (blue numbers indicate waves). The bottom panel plots smoothed local *R*-estimates (red with 95% confidence bands) and associated *Z* numbers (blue) measuring the % likelihood of elimination.

Initially, we infer a largely supercritical *R* that appears to fall swiftly in response to emergency NPIs that were engaged without delay. This potentially minimised the size of wave 1 (subject to diagnostic testing rates). Subcritical transmission followed across wave 2, making the strategic border closures apt and effective and suggesting that this wave was not allowed to become a genuine resurgence. Intriguingly, soon after *Z* starts to increase, achieving 55% in early May. Genetic data indicate that between waves 2 and 3 there was elimination of one circulating strain of SARS-CoV-2 [personal communication, B Cowling]. The peaking of our *Z* number might reflect this, with co-circulating lineages preventing complete viral elimination. The weak transmission we infer in May-June 2020 supports the NPI relaxation that occurred. Interestingly, we find no evidence of increasing elimination potential in July.

This belies what might be naively expected, given that zero local cases were recorded for 21 consecutive days, which is 7 days below the WHO elimination criterion. However, on that 21^st^ day we infer *Z* ≈ 0%, emphasising the utility of end-of-epidemic metrics that consider the local-import transmission context [15]. After this decrease in *Z*, our framework confidently signals supercritical community transmission, which likely created a large wave 3. The steep rise in *R*-estimates is already underway by that 21^st^ 0-case day, showing how maximally informed outbreak analytics can help decipher transmission signals, which are unclear from case-data. We infer change-points in *R* that correlate with timing of key NPIs and find that those measures eventually constrict COVID-19 spread (*R* < 1 in August). As wave 3 wanes we obtain evidence supporting the NPI release from September (*R* declines) but then flag another confident rise in transmission in late November. This coincides well with the wave 4 declaration.

### Resurgence and eventual elimination in Victoria state, Australia

Australia reported its first cases of community transmission on March 2, 2020. Victoria state, where many cases were beginning to concentrate, declared a state of emergency on March 16, which included stay at home orders and many activity restrictions. Further, all Australian borders were closed on March 20. This likely reduced both community-spread and capped the influence of imported cases in April, minimising the initial wave. As cases declined NPIs were adaptively relaxed and re-introduced across May and June. While a large case-cluster was discovered across May 2-14, linked to a meat packing plant and contributing the majority of infections in that time-period, swift contact-tracing and quarantines contained its impact [24]. However, the relaxation of household mixing NPIs resulted in large household gatherings that led to a rise in local cases. Victoria responded with postcode-based lockdowns by 30 June.

However, this was not sufficient and local cases burgeoned. NPIs were ramped up throughout July but the second wave grew exponentially. A 4-stage restriction policy with a target of zero community transmission was enacted and reinforced, building to a major lockdown (stage 4) on August 2 [24]. A state of disaster was also declared. Restrictions included confinement at home, curfews and closures. Slowly this large wave subsided and over 100 days after, NPIs began being relaxed from October 18 (stage 3). Staged re-opening continued until November 22 (stage 1), when most restrictions were removed. By November 27, the last time-point we consider, Victoria state had recorded 28 days of 0-cases and declared elimination of SARS-CoV-2. This timeline and the epidemic curve are given in Figure 3 (top), with data from [25]. We now investigate the transmission dynamics underlying this data using our *R-Z* framework.

**Figure 3:**
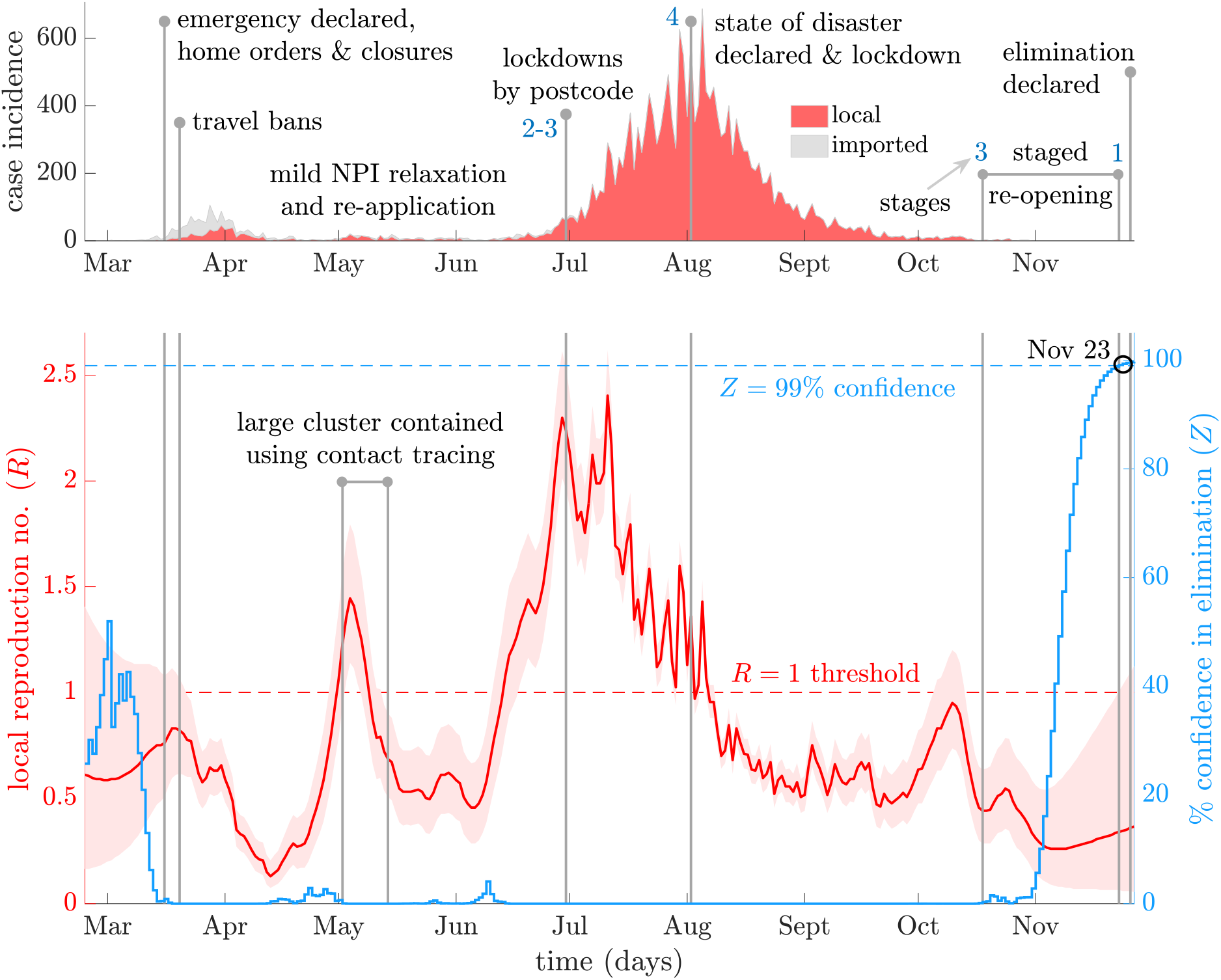
Local transmission dynamics of COVID-19 in Victoria, Australia. The top panel illustrates local (red) and imported (grey, stacked) cases by diagnosis date from [25]. Vertical lines highlight important policy change-times and responses (blue numbers are NPI restriction stages). The bottom panel presents smoothed local *R*-estimates (red with 95% confidence bands) and resulting *Z* numbers (blue) measuring the % probability of elimination.

Our main results are displayed in Figure 3 (bottom). We find a slow start to the initial wave in Victoria with the confidence region of *R* only partially above 1, *Z* at moderate levels and most cases being imported. However, *Z* = 0% quickly occurs and *R* begins to increase. The speedy declaration of emergency and travel ban precede a clear downward trend in our *R*-estimates associated with suppression of this wave. This could have been especially effective since the majority of cases then were imported. Following this local *R* remains subcritical, corroborating Victoria’s adaptive NPI relaxation. We observe a large swing in our *R*-estimates that roughly aligns with the meat-packing plant cluster. The rapid peak and fall in *R* likely reflect the contact-tracing employed on this single cluster, which forms most cases in this period. Consequently, no true resurgence occurs, until a month later when our *R*-estimates rapidly elevate.

This provides warning of the explosive second wave. The much larger local *R* observed here suggests this period was the most critical for COVID-19 transmission in Victoria. Steadily harsher NPIs (stages 2 to 4) are supported and correspond to *R* falling below 1. This fall is slower than its initial rise, expressing how larger-sized epidemics can be difficult to control and evidencing the need for sustained lockdown. Fluctuations about this *R* trend in July-August reflect weekend biases in reporting (likely exacerbated by large case counts) and disappear if the incidence is first treated with a 7-day moving average filter (see Supplement Figure J). Transmission remains subcritical for most of September and October. The stifled community spread corroborates the staged reopening strategy. As cases continually fall, local *Z* increases also supporting NPI release. We obtain *Z* ≈ 99% by November 23, 2020, which favours NPI relaxation (stage 1), bolsters evidence for the success of Victoria’s elimination-based strategy and suggests that we have almost 100% confidence in the official end-of-epidemic declaration.

## Discussion

Understanding the transmission forces underpinning epidemic elimination and resurgence is critical to the efficient design and timely implementation of NPIs. Appropriate responses to import-driven versus locally sustained outbreaks can differ markedly and materially given the constraints on resources. While naïve *R*-estimates and cross-country comparisons have been popularised across the COVID-19 pandemic, we argue that locally relevant strategies tuned to the specific dynamics of an area are imperative. Our proposed early-warning *R*-*Z* framework can support this aim, especially in the crucial data-limited lull between potential epidemic waves, where it significantly improves our ability to reliably denoise transmission change-point signals and decipher indicators of upcoming epidemic dynamics [16].

We attained this improvement by harnessing methodology from signal processing and control engineering [17]. Common *R*-estimators only exploit some of the information encoded within incidence data. Our Markovian smoothed *R* applies forward-backward algorithms aimed at maximum information extraction (see Methods). This can double statistical efficiency in some instances [19]. Combining this methodology with local-import models [13], we derived a local *R*-estimator that is robust at small incidence and identifies change-points naturally. Reliable change-point detection can be problematic for existing estimators [9], while limited robustness hinders inference of elimination likelihoods [15]. Our *R* (Eq.1) allowed us to devise *Z* (Eq.2), a new metric for ascertaining the confidence in elimination that overcomes this issue.

We showcased our *R-Z* framework on important and diverse COVID-19 case-studies (see Results). New Zealand, Hong Kong and Victoria state have presented difficulties to standard analyses due to prolonged low-incidence durations and large imported case numbers [1,3–6]. Although the categorisation and types of NPIs used differ, our analyses present clear evidence for the effectiveness and timeliness of the strategies employed in all three regions of study (Table 1). We inferred sharp correlations between downward transitions in *R* and the timing of key NPIs, with major *R* reductions seen after 2 weeks of sustained NPI usage. We estimated that the swiftness of NPI enforcements in several instances, such as New Zealand’s second wave and Hong Kong’s first wave, may have averted more explosive resurgence, as illustrated by clear *R* turning points. We also found that NPIs were often sustained until local transmission was suppressed, supporting the choice of their relaxation or release.

**Table 1:** Alignment of NPIs with inferred *R-Z* metrics. We summarise how the timing of key NPI applications and relaxations as well as official declarations of elimination correlate with salient transmission dynamics, as estimated under our *R-Z* framework for COVID-19 in New Zealand, Hong Kong, China and Victoria state, Australia, across 2020 (Figures 1-3).

| New Zealand | Policy actions and details | Early-warning $R$-$Z$ evidence |
| --- | --- | --- |
| Mar 19-26 | Border closures, nationwide lockdown and alert level 4. | $R = 1.32$ (0.94, 1.78) at action-point. Falls to 1.18 (0.97, 1.41) after 1 week and then 0.68 (0.56, 0.82) at 2 weeks. |
| May 14-Jun 9 | Relaxation of some controls, de-escalation to alert level 2. | At beginning and end of period $R = 0.38$ (0.17, 0.70) and 0.25 (0.06, 0.64). In period it is comfortably below 1. |
| Jun 9 | End-of-epidemic declaration (WHO criteria – 28 days of no cases), alert level 1. | $Z = 98.7\%$ at declaration. From Jun 5-15 it rises from 96.1% to 99.8%. A declaration within this time is at least 95% certain. Rapid fall to $Z = 26.7\%$ on Jun 16. |
| Aug 12 | Work at home, closures and bubbles. Alert levels 2-3. | At action-point $R = 1.80$ (1.36, 2.31). Falls to 1.15 (0.88, 1.47) 1 week later and 0.75 (0.54, 1.00) after 2 weeks. |

| Hong Kong | Policy actions and details | Early-warning $R$-$Z$ evidence |
| --- | --- | --- |
| Jan 25 | Wave 1, state of emergency declared, NPIs enforced e.g., schools kept closed | $R = 1.38$ (0.80, 2.14) at action-point. Falls to 1.17 (0.80, 1.63) after 1 week and then 0.75 (0.49, 1.07) at 2 weeks. |
| Mar 25-29 | Wave 2, border closures and bans on public gatherings. | $R = 0.41$ (0.31, 0.54) on Mar 25. Reduces to 0.12 (0.07, 0.20) and then 0.06 (0.02, 0.13) at 1 and 2 weeks after Mar 29. |
| May 4, Jul 5 | Extinction of a SARS-CoV-2 lineage (in May), 21 days of no local cases Jul 5). | $Z = 54.9\%$ on May 4 and on average 47.2% across Apr 29-May 6. Partial possibility of elimination in this period. $Z = 0.02\%$ on Jul 5 – effectively no chance of elimination. |
| May 5-27 | NPIs gradually relaxed. Lull between potential waves. | At beginning and end of period $R = 0.20$ (0.07, 0.45) and 0.29 (0.12, 0.54). In period it is comfortably below 1. |
| Jul 13-19 | Wave 3, work-from-home orders, venue closures and social distancing measures. | $R = 1.76$ (1.53, 2.00) at first action-point. Falls to 1.15 (1.02, 1.30) and 0.62 (0.52, 0.72) 1 and 2 weeks after Jul 19. |
| Sept 1-Nov 1 | Gradual relaxation of NPIs | At beginning and end of period $R = 0.50$ (0.37, 0.67) and 0.30 (0.18, 0.48) and never above 1 throughout. |
| Nov 23 | Wave 4, NPIs re-imposed. Last point analysed. | $R = 1.27$ (0.99, 1.59) at action-point. Early-warning of wave of local transmission. |
| Victoria state | Policy actions and details | Early-warning $R$-$Z$ evidence |
| Mar 16-20 | Stay-at-home orders, state of emergency declared. Added national travel bans. | $R$ rises from 0.76 (0.50, 1.08) to 0.82 (0.61, 1.06) across period. After 1-2 weeks it is 0.59 (0.48, 0.74) and 0.48 (0.39, 0.60). |
| May 2-14 | Large cluster of cases from meat packing plant. Contact tracing and isolation applied. | At beginning of period $R = 1.22$ (0.93, 1.57). It falls to 0.68 (0.50, 0.90) by the end. NPIs seem effective. |
| Jun 30 | Adaptive lockdowns, stage 2 and 3 restrictions engaged. | $R = 2.24$ (1.96, 2.53) at point applied. Falls to 2.12 (1.91, 2.35) and 1.67 (1.52, 1.83) after 1-2 weeks. Further NPIs needed. |
| Aug 2 | Complete lockdown, stage 4 restrictions. State of disaster officially declared. | $R = 1.35$ (1.26, 1.45) at action-point. Might be larger due to noise in cases here. Falls |
|  |  | to 0.81 (0.74, 0.88) and 0.71 (0.64, 0.78) at 1 and 2 weeks after imposition. |
| Oct 18-Nov 22 | Gradual NPI release, from stages 3 to 1. | At beginning and end of period $R = 0.44$ (0.28, 0.63) and 0.33 (0.07, 0.97). In period it remains safely below 1. |
| Nov 27 | End-of-epidemic declaration (WHO criteria – 28 days of no cases), alert level 1. | $Z = 99.7\%$ at declaration. From Nov 17-27 it rises from 95.0% to 99.1%. A declaration within this period is at least 95% certain. |

However, while effective, the responses of our study regions were not all perfect. We observed right skew in the dynamics of *R* – its growth was generally faster than its decay – accentuating the need for rapid NPI application. Hong Kong’s third wave and Victoria’s second wave both had notable periods over which *R* climbed steadily above 1. Imposing NPIs 1-2 weeks earlier might have appreciably reduced the epidemic burden in these cases. Stricter handling of imported cases may also be important going forward, as several subsequent waves were kickstarted by repeated introductions. Hong Kong’s second wave and Victoria’s first wave, which featured early travel bans and closures that suppressed the influence of many imported cases, might serve as a good template for handling such scenarios.

New Zealand and Victoria state both initiated and successfully implemented elimination-based strategies. We inferred *Z* ≈ 99% confidence in the end-of-epidemic declarations made by both regions, rigorously backing those decisions. We estimated that Hong Kong attained Z ≈ 50-55% in synchrony with the believed extinction of one circulating SARS-CoV-2 strain. Co-circulating lineages may have prevented the achievement of elimination. We observed *Z* ≈ 0% despite sustained *R* < 1 in many periods, highlighting the insufficiency of *R* for assessing elimination. Maintaining NPIs until *Z* crosses some threshold could be one data-informed way of deciding when to safely relax measures. Overall, we conclude that all regions responded decisively and adaptively to fluctuating local transmission. This conclusion is not well-supported by more naïve *R*-estimates that neglect local-import models or non-smoothed ones that fail to use all the information in the incidence data (see Supplement for these analyses).

While our results provide rigorous underpinning and insight into COVID-19 dynamics in New Zealand, Hong Kong and Victoria, there are limitations. We do not explicitly compensate for reporting delays or under-reporting. However, these issues are likely minimised by the high fidelity of surveillance, contact-tracing and testing in our case-studies. Hong Kong had rapid screening systems ready due to past experiences with SARS in 2003 [22], while aggressive testing strategies in New Zealand and Australia have garnered praise [24]. Delays from symptom-onset to case notification in New Zealand are just 1.7 days, for example [20]. Moreover, we obtained strong one-step-ahead predictive fits (see Methods and Supplement), indicating model adequacy [9]. We also do not factor in time-varying serial intervals [26] or asymptomatic spread. However, limited data on these preclude improvement of our estimates. If, for some location of interest, surveillance biases are known to be significant and relevant data are available (e.g., on reporting and serial interval fluctuations), then we recommend first compensating for these biases to derive the best possible incidence curve, and then applying our framework. This can be achieved by pre-processing the reported incidence to minimise the influence of these biases. For example, weekend surveillance effects can be corrected with weekly case averaging (see Supplement Figure J) and estimates of time-varying reporting fractions, if available, can be used to up-sample known cases to gauge true incidence [27]. Additional noise sources may be modelled by generalising our Poisson observation model (see Methods) to include further dispersion (e.g., negative binomial descriptions). Auxiliary data sources, such as genomic sequences, can also be used to derive independent incidence curves [28], which may be input into our *R-Z* method to improve the reliability of inferred trends.

Delays or latencies in data collection can be resolved via deconvolution algorithms or simple mean shifting provided information on those delays are available [29], while up-to-date serial intervals derived from contact-tracing or other surveillance can be incorporated directly within our methodology. Under-ascertainment, asymptomatic spread and problems stemming from approximating the generation time by the serial interval (see Methods) are more difficult to correct but solutions are actively being researched [30]. Note that early-warning signals are fundamentally not possible if outbreak monitoring is poor (e.g., if there are large latencies to case notification). We envision our framework as supplementing outbreak analytics toolkits of regions with dedicated surveillance programmes.

Our *R-Z* framework is available in the *EpiFilter* package https://github.com/kpzoo/EpiFilter as a major extension. Although we analysed countries and regions, we expect our methodology to be particularly useful at finer scales, where incidence is necessarily smaller by division. There, reliable signalling of transmission change-points might support more targeted and less disruptive NPIs (e.g., postcode-lockdowns versus nationwide ones). Our method only requires clear classification of local and imported cases to remain valid, is reproducible and easy to run with minimal computational overhead. Early and robust warnings of resurgence or elimination can distinguish timely from tardy interventions. Local and contextualised metrics, such as *R-Z*, will hopefully help separate the signal from the noise, when it comes to effective NPIs.

## Methods

The renewal transmission model is a popular and flexible means of modelling the spread of an infectious disease [31]. It describes how the number of new cases, i.e., the incidence, at time *s*, denoted I_s_, depends on the effective reproduction number at that time, R_s_, and the past incidence, which is summarised by the total infectiousness, Λ_s_, as follows.

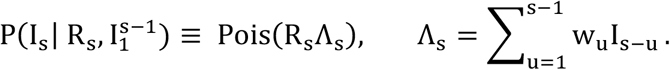

Here Pois indicates Poisson, ≡ denotes equality in distribution and *w*_u_ is the probability that it takes *u* days between the time of infection of a primary and secondary case. We consider incidence curves observed over times 1 ≤ s ≤ t with 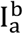 as the portion {I_a_, I_a+1_, …, I_b_}.

The *w*_*u*_ for all *u* define the generation time distribution of the infectious disease of interest. We make the standard assumption that this distribution is known and well approximated by the serial interval distribution [7]. For the SARS-CoV-2 virus we use Gam(2.37, 2.74) [32], which has a mean of 6.5 days. However, we find our key results are robust to other estimated SARS-CoV-2 serial interval distributions [33]. While we do not account for possible changes to the serial interval distribution (e.g., contractions due to NPIs [26]) or for temporal variations in case ascertainment [15], this model remains valid if those changes are known and included [7].

Since time-varying reproduction numbers are likely to be autocorrelated [34], we generalise the renewal model to include a minimal, Markov random-walk assumption. The subsequent dynamical state model results, with Norm representing normally distributed noise.

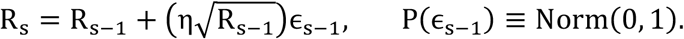

Here *η* is a state noise parameter, which is easy to tune and set to 0.1 in all analyses [16]. We validate this choice using cumulative one-step-ahead log-predictive fits, which show that, at this *η*, we predictively and sequentially reproduce the observed incidence in each case study with minimal generalisation error (see Supplement Figures B, E and H). More details on this information theoretic model adequacy test, known as the accumulated predictive error (APE) metric, are provided in [9,35]. Our state noise also models some heterogeneous transmission (because it leads to a doubly stochastic Poisson description of incidence), which is a salient characteristic of many infectious diseases, including COVID-19 [34,36].

Our description offers two main advantages. First, we do not need to specify predetermined change-points or averaging windows as in many popular approaches (e.g. *EpiEstim* [7,37]). Inference of *R* is known to be highly sensitive to window-size and change-times choices [8,9]. Second, because we only make minimal state-assumptions, our estimates are less controlled by prior model assumptions [38]. Using these equations as is, however, only yields naïve *R*-estimates, as no distinction has yet been made between local and imported cases.

To incorporate case introductions, we apply a key decomposition [13]. If L_s_ is the number of local cases at *s* and M_s_ the imported ones, then I_s_ = L_s_ + M_s_. Both types of cases drive future local infections and so the transmission model is extended [18].

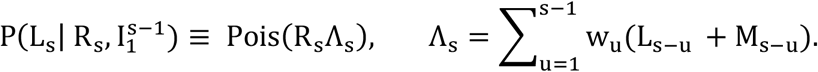

The state model is unchanged but now describes the evolution of local reproduction numbers. We next outline how to obtain *R*-estimates from the above transmission and state models that improve the robustness and reliability of inference when incidence is small.

We can construct three possible posterior distributions to describe how information from an observed incidence time-series is recruited to form estimates of R_s_. These are known as the filtering (**P**_**s**_), predictive (**r**_**s**_) and smoothing (**q**_**s**_) posterior distributions.

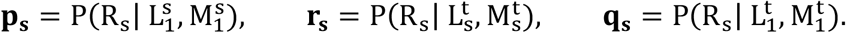

These distributions are fundamental to any real-time or retrospective estimation problem and have been studied deeply in control systems engineering and signal processing [17,39].

Standard inference methods either approximate **P**_**s**_ (e.g., *EpiEstim*) or **r**_**s**_ (e.g., the *Wallinga-Teunis method* [10]), which respectively incorporate past (backward) or “future” (forward) incidence information. In both instances, estimates suffer from edge-effects [7] and are more vulnerable to low-incidence periods because they cannot exploit all the available information [16,19]. This can be a significant limitation, especially in the important lull between potential epidemic waves, where data are scarce, but reliable estimates are vital for preparedness. A key contribution of this study is the computing of **q**_**s**_ to derive smoothed, local *R*-estimates that formally utilise all incidence information up to s [17], under our generalised local-import model.

We achieve this by adapting recursive forward-backward Bayesian algorithms from [16]. First, we iteratively calculate **P**_**s**_ (the forward pass) as below.

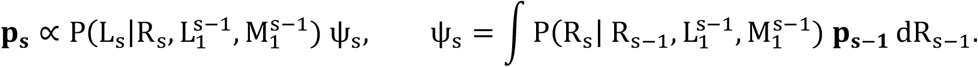

We apply a uniform prior distribution for **P**_**1**_. The **P**_**s**_ distributions are fed-back successively to then obtain **q**_**s**_ (the backward pass) resulting in Eq.1, with **q**_**t**_ = **P**_**t**_

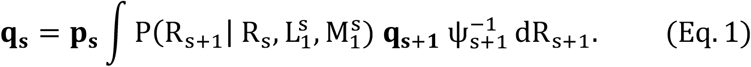

By constructing **q**_**s**_ we maximise the information that is integrated into our local transmission estimates and minimise undue dependence on prior model choices and assumptions [9,38].

This upgrades overall performance, when compared to common *R*-estimation methods, and significantly bolsters estimate robustness when incidence (or related) data are limited. These improvements follow because **q**_**s**_ ≈ ∝ **r**_**s**_ **p**_**s**_ P(R_s_)^−1^ [16], meaning that it explicitly integrates information from both **P**_**1**_ and **r**_**1**_ Further details on these recursive algorithms are provided in [17,39], where it is also noted that this formulation results in *R*-estimates that minimise mean squared estimation errors (relative to their “true” values). These estimates can be updated in real time as more data accumulate (time *t* increases).

Our novel local *R*-estimates are functions of **q**_**s**_ (for example our mean estimate is ∫ **q**_**s**_ R_s_d *R*_*s*_) that can decipher important early-warning signals of upcoming resurgence (see Results and Supplement) often buried in low-incidence data. However, this is not sufficient to assess the chance of local elimination. We therefore introduce the local, smoothed *Z* number, a new measure of the statistical lifetime of the epidemic, which is obtained by generalising the recent theory from [15] to incorporate the **q**_**s**_ distribution from the import-local transmission model.

We define our % confidence in an epidemic being eliminated (i.e., propagating no future local cases) at time *s* as 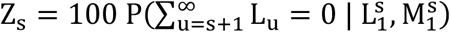 with 100 – Z_s_ as the survival likelihood of at least one future case given available data. We can solve for Z_s_ by appending a pseudo stream of 0-incidence values 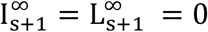 and deriving posterior distributions over 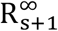 assuming these 0-data. Expanding *Z*_*s*_ sequentially we get the product below [15].

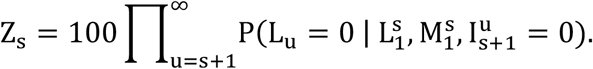

Using the above local-import renewal model each term can be obtained as a function of *R*_*u*_. Next, we marginalise over our *R*-distributions of interest, assuming the pseudo-data to obtain Eq.2, which together with Eq.1 forms our *R-Z* framework.

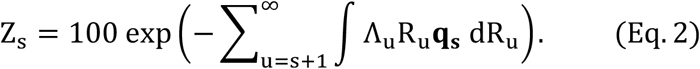

This **q**_**s**_ is obtained from the same smoothing algorithms above but under the assumption that there are no future local cases. It is accordingly recomputed in real time for every *s*. We can also substitute **q**_**s**_ with either **p**_**s**_ or **r**_**s**_ to obtain related elimination metrics.

Our *Z* number quantifies the confidence that a local epidemic is over, given past incidence and in the presence of imported cases. This measure is more adaptive and telling than current WHO guidelines, which propose end-of-epidemic declarations based on fixed waiting times that relate to twice the incubation period of the infectious disease [12,15]. It also appreciably improves on previous metrics proposed in [15], which were limited by the destabilisation of *R* at low incidence and unable to incorporate the uncertainty in local transmissibility.

Thus, our *R-Z* framework makes minimal assumptions and can be applied in real-time to infer the upcoming risk of local transmission or the emerging likelihood of elimination. It can also be used retrospectively to discriminate among hypotheses surrounding the effectiveness and timeliness of previously implemented NPIs. Importantly, it remains valid at many scales of interest, e.g., countries or sub-regions of a state, provided local and imported case data are delineated. In this development we have assumed the availability of high-quality incidence data. If significant reporting delays and biases exist, these should be first compensated for (see Discussion for details) before applying our framework.

We showcase the power of this framework (both in real time and retrospectively) on three COVID-19 case-studies in the Results and Supplement. These examples all feature prolonged low-incidence periods that have destabilised standard inference methods [1,3–6]. Hopefully, our framework, by minimising the noise and maximising the informativeness of estimates, will help to better target and time NPI application and relaxation for a given region of interest. It is available as part of the *EpiFilter* package at: https://github.com/kpzoo/EpiFilter.

## Supporting information

supplementary analyses

## Data Availability

The computational framework introduced in this study forms a major update of the EpiFilter codebase, which is available in R and Matlab (at branches matching case study names) at: https://github.com/kpzoo/EpiFilter

## Authors Contributions

KVP and CAD conceptualised the study. KVP performed the research, developed software, methodology and wrote the initial draft. All authors validated results and wrote the final draft.

## Data Accessibility

Code to reproduce the analyses in this paper is available as a major update to the EpiFilter package [16] on alternate branches of https://github.com/kpzoo/EpiFilter.

## Funding

KVP and CAD acknowledge joint centre funding from the UK Medical Research Council (MRC) and Department for International Development (DFID) under grant reference MR/R015600/1. CAD thanks the UK National Institute for Health Research Health Protection Research Unit (NIHR HPRU) in Emerging and Zoonotic Infections in partnership with Public Health England (PHE) for funding (grant HPRU200907). The funders had no role in study design, data collection and analysis, decision to publish, or preparation of the manuscript.

## Competing interests

The authors declare no competing interests.

## Notes

### Competing Interest Statement

The authors have declared no competing interest.

### Summary of Updates

Additional analyses included in Supplement and text updated in the Main Document.

