## supplementary analyses for "Deciphering early-warning signals of SARS-CoV-2 elimination and resurgence from limited data at multiple scales"

### New Zealand: real-time *R*-estimate comparisons

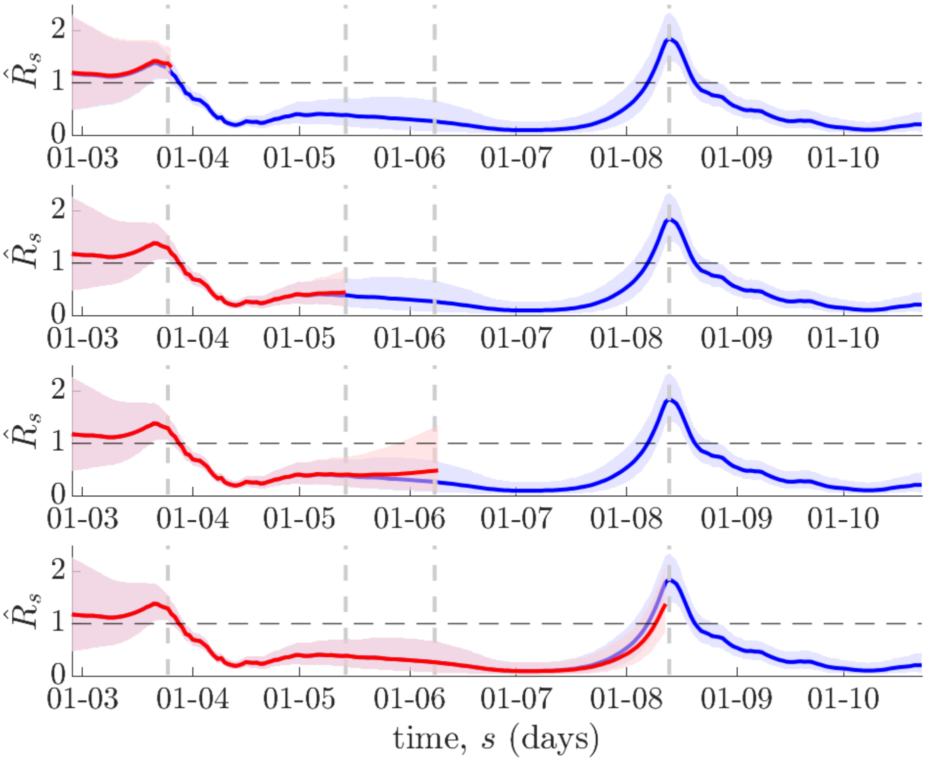

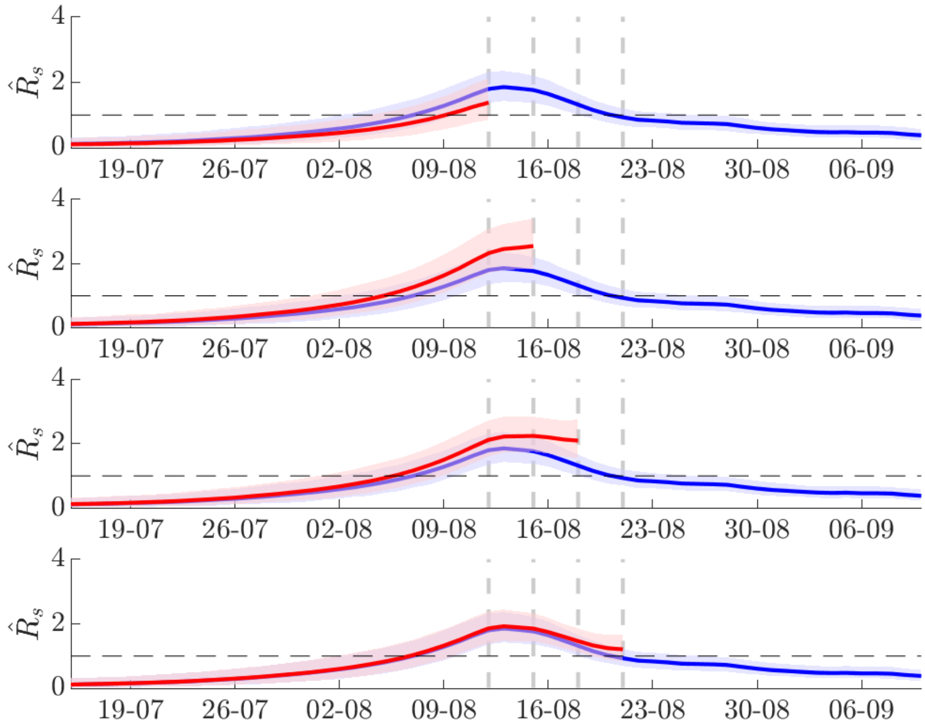

**Figure A: Real time *R*-estimates for New Zealand**. (Top) Grey lines indicate the times of key policy actions that correspond to Figure 1 of the main text. Each panel examines the smoothed, local *R*-estimates (see Methods of the main text) computed from data up to each policy action time (red with 95% confidence bands) as compared to those from Figure 1 (blue with 95% confidence bands), which processes the entire incidence curve. There is a good correspondence between the real time and retrospective estimates indicating the ability of the framework to provide early-warning signals. (Bottom) The main discrepancies are highlighted. Grey lines indicate successive 3-day releases of data. Most discrepancies (and particularly signals of resurgence) are largely resolved by the first 3-day release of data.

**New Zealand: smoothed vs standard (local) *R*-estimates**

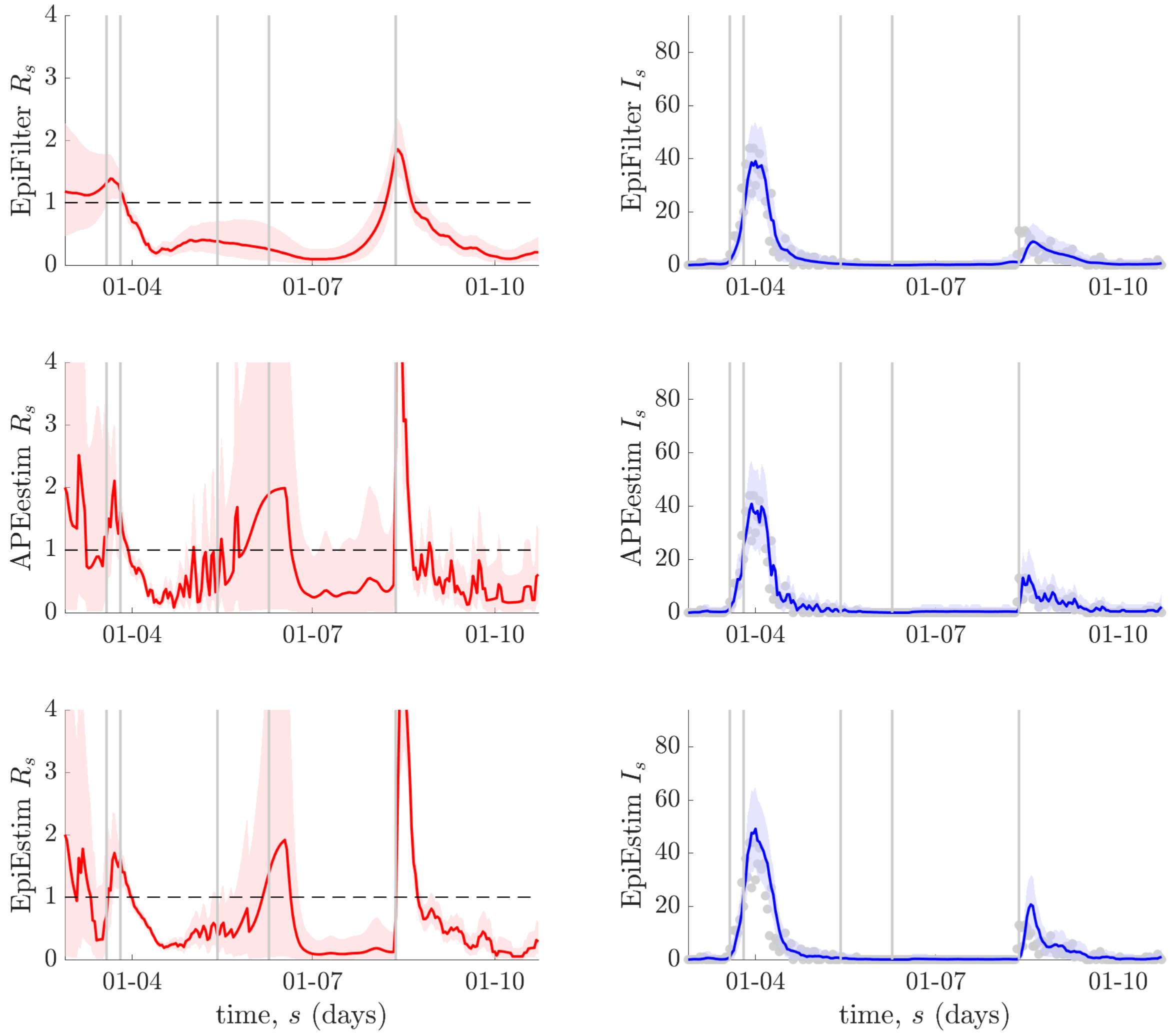

**Figure B: Local *R*-estimates for New Zealand**. Grey lines indicate the times of key policy actions as shown in Figure 1 of the main text. Left panels illustrate different *R*-estimates (red with 95% confidence bands) using *EpiFilter* (top panels, see Methods of main text), *APEestim* (middle panels, optimises window length for prediction) and *EpiEstim* (bottom panels, uses a weekly window length). Right panels provide one-step-ahead predictive fits (blue with 95%

confidence bands) of the actual incidence data (grey circles) that result from the *R*-estimates of the corresponding left panel. Both *APEestim* and *EpiEstim* only use backward information, while *EpiFilter* exploits both backward and forward information, resulting in better stability and reliability, especially when incidence is small.

#### New Zealand: local vs naïve (smoothed) *R*-estimates

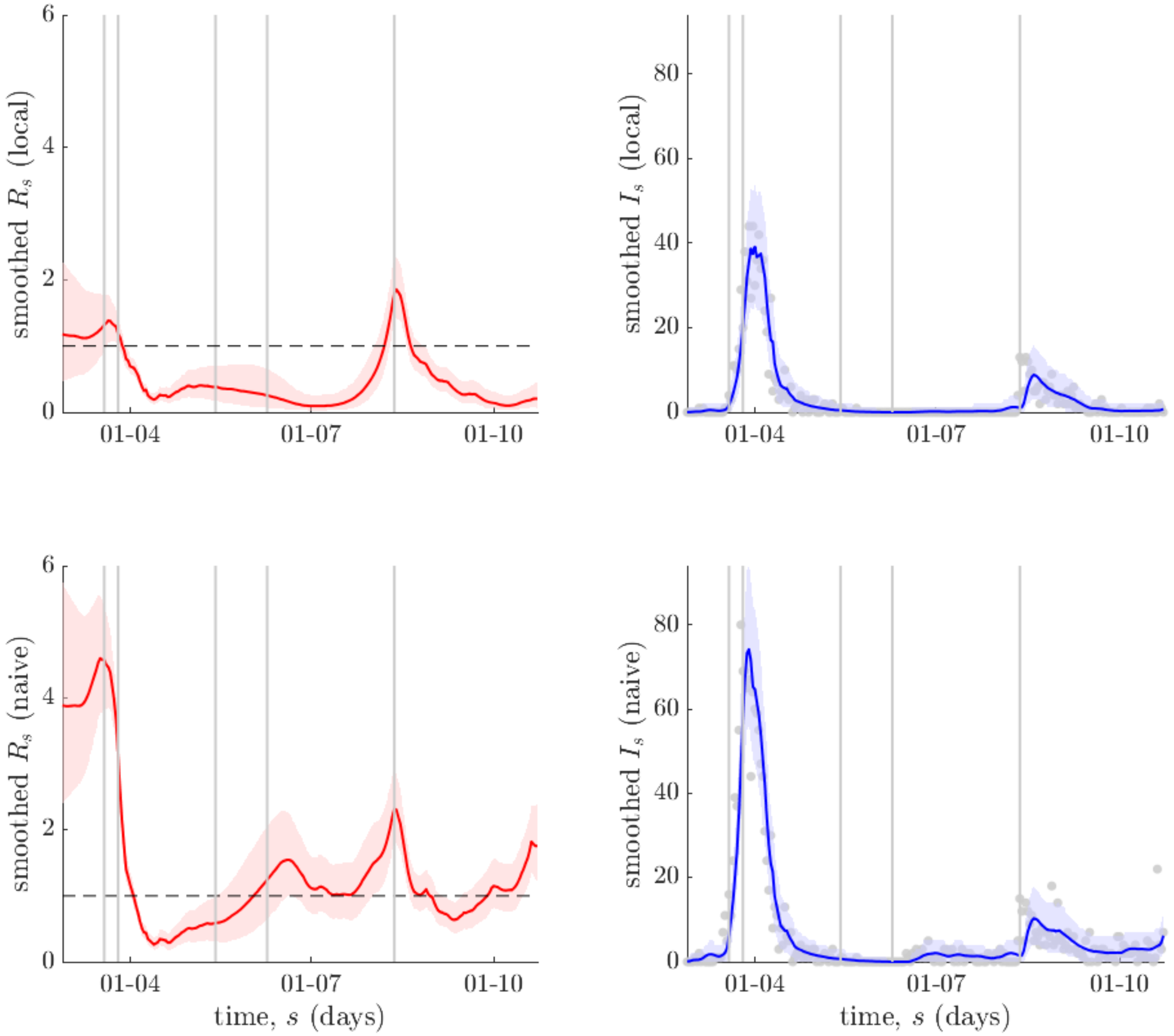

**Figure C: Smoothed *R*-estimates for New Zealand**. Grey lines indicate the times of key policy actions, corresponding to Figure 1 of the main text. Left panels illustrate the difference in smoothed *R* estimates (red with 95% confidence bands) (i.e., they use both forward and backward incidence data – see Methods of the main text and the *EpiFilter* package) using renewal models that account for (local, top panels) or ignore (naïve, bottom panels) the differing impact of local and imported cases. Naïve estimates significantly over-estimate *R*. Right panels provide one-step-ahead predictive fits of the incidence (blue with 95% confidence bands) that result from these *R*-estimates. The actual incidence data are grey circles. The fits suggest that the data are well represented by the model.

**Hong Kong: real-time *R*-estimate comparisons**

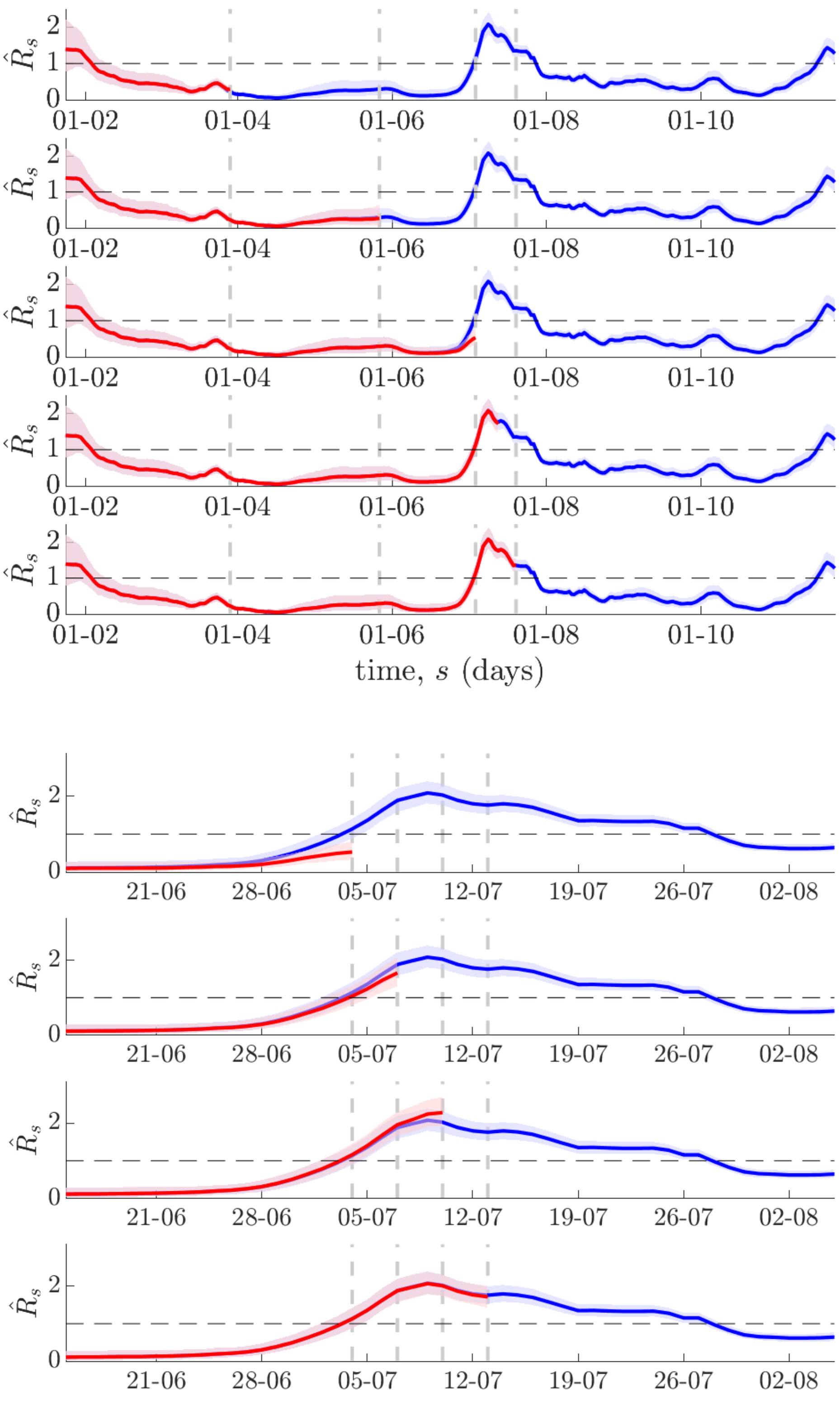

**Figure D: Real time *R*-estimates for Hong Kong**. (Top) Grey lines indicate the times of key policy actions that correspond to Figure 2 of the main text. Each panel examines the smoothed, local *R*-estimates (see Methods of the main text) computed from data up to each policy action time (red with 95% confidence bands) as compared to those from Figure 2 (blue with 95% confidence bands), which processes the entire incidence curve. There is a good correspondence between the real time and retrospective estimates indicating the ability of the framework to provide early-warning signals. (Bottom) The main discrepancies are highlighted. Grey lines indicate successive 3-day releases of data. Most discrepancies (and particularly signals of resurgence) are largely resolved by the first 3-day release of data.

#### Hong Kong: smoothed vs standard (local) *R*-estimates

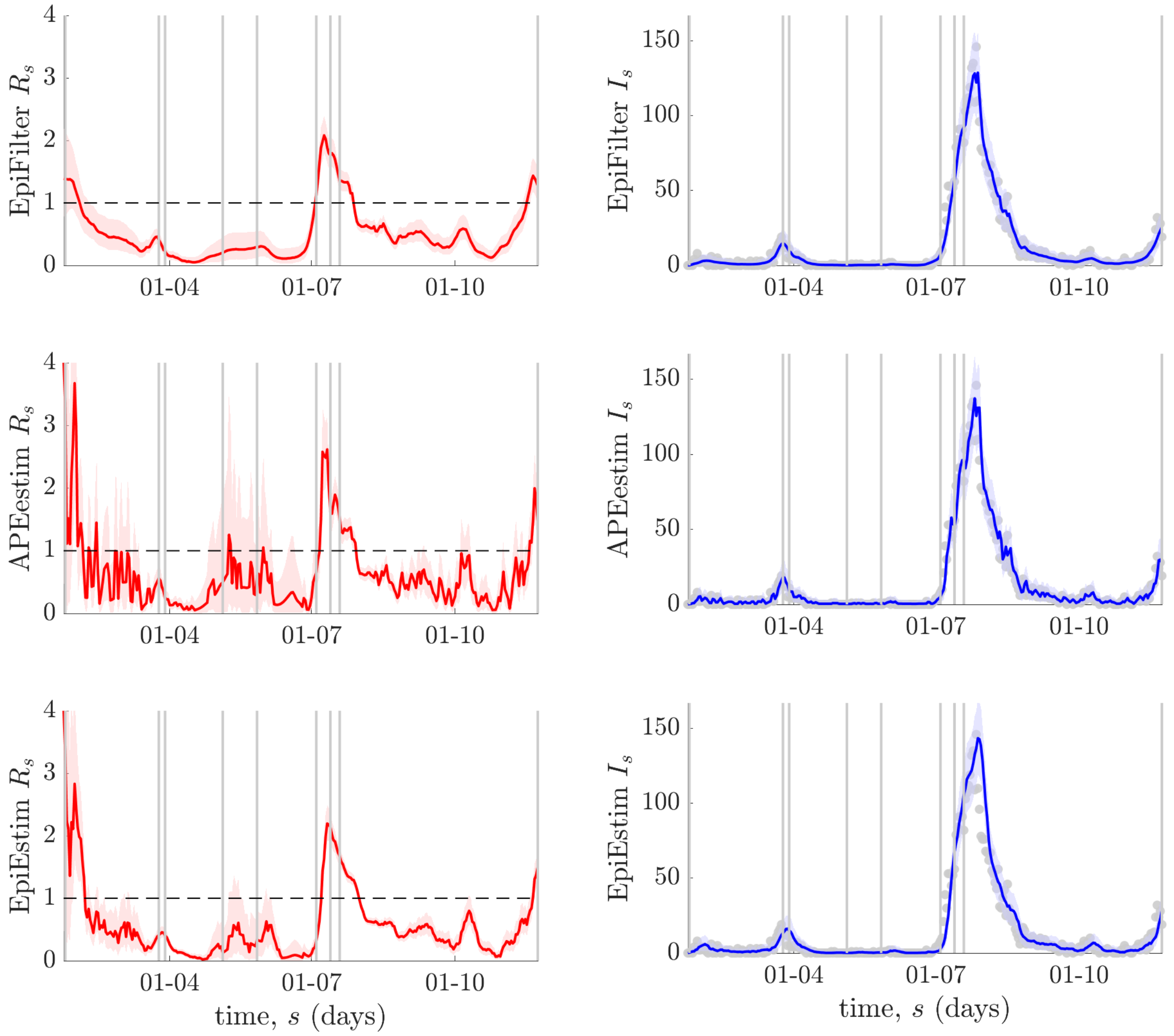

**Figure E: Local *R*-estimates for Hong Kong, China**. Grey lines indicate the times of key policy actions as shown in Figure 2 of the main text. Left panels illustrate different *R*-estimates (red with 95% confidence bands) using *EpiFilter* (top panels, see Methods of main text), *APEestim* (middle panels, optimises window length for prediction) and *EpiEstim* (bottom panels, uses a weekly window length). Right panels provide one-step-ahead predictive fits (blue with 95% confidence bands) of the actual incidence data (grey circles)

that result from the *R*-estimates of the corresponding left panel. Both *APEestim* and *EpiEstim* only use backward information, while *EpiFilter* exploits both backward and forward information, resulting in better stability and reliability, especially when incidence is small.

#### Hong Kong: local vs naïve (smoothed) *R*-estimates

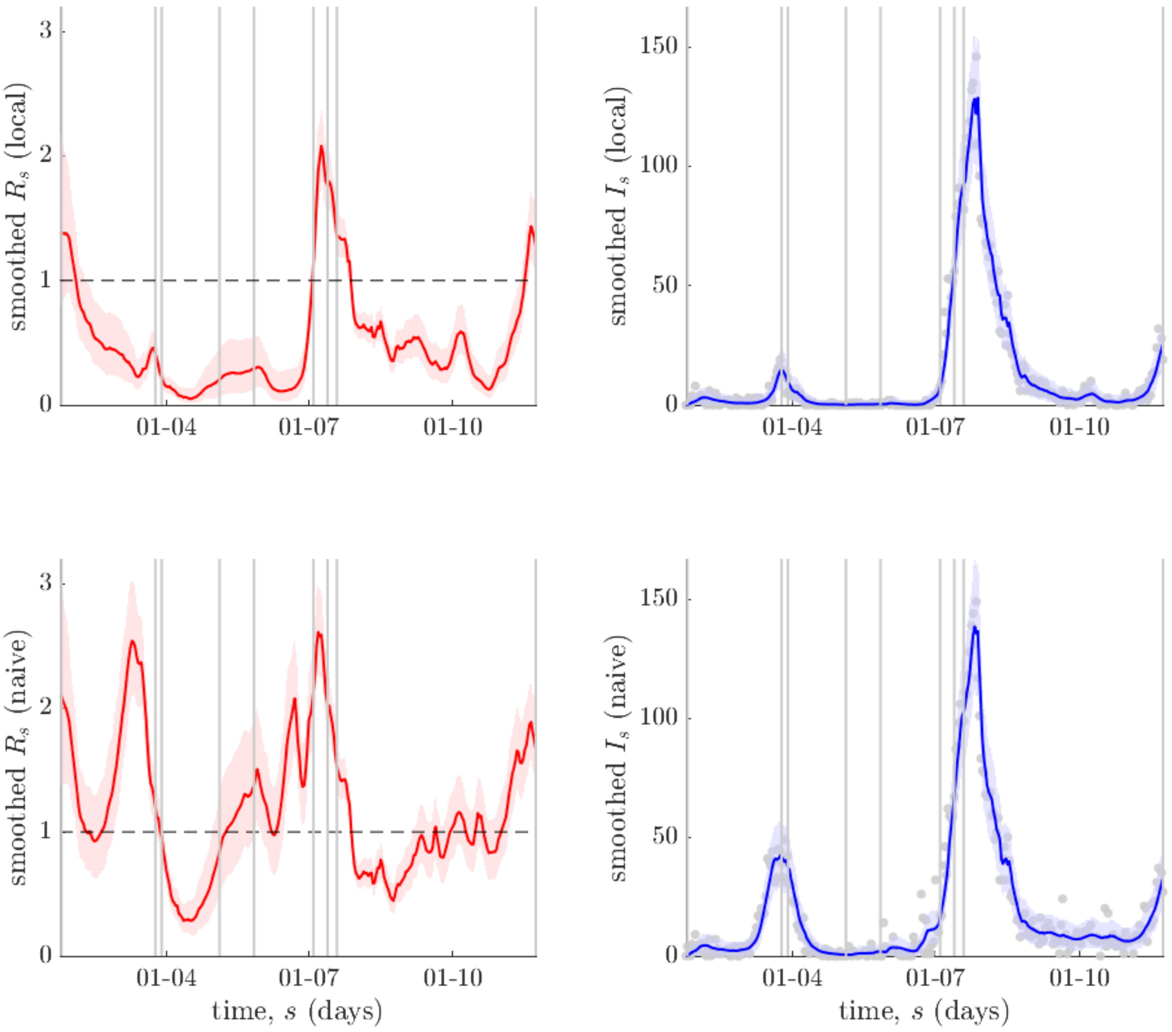

**Figure F: Smoothed *R*-estimates for Hong Kong, China**. Grey lines indicate the times of key policy actions, corresponding to Figure 2 of the main text. Left panels illustrate the difference in smoothed *R* estimates (red with 95% confidence bands) (i.e., they use both forward and backward incidence data – see Methods of the main text and the *EpiFilter* package) using renewal models that account for (local, top panels) or ignore (naïve, bottom panels) the differing impact of local and imported cases. Naïve estimates significantly over-estimate *R*. Right panels provide one-step-ahead predictive fits of the incidence (blue with 95% confidence bands) that result from these *R*-estimates. The actual incidence data are grey circles. The fits suggest that the data are well represented by the model.

#### Victoria: real-time *R*-estimate comparisons

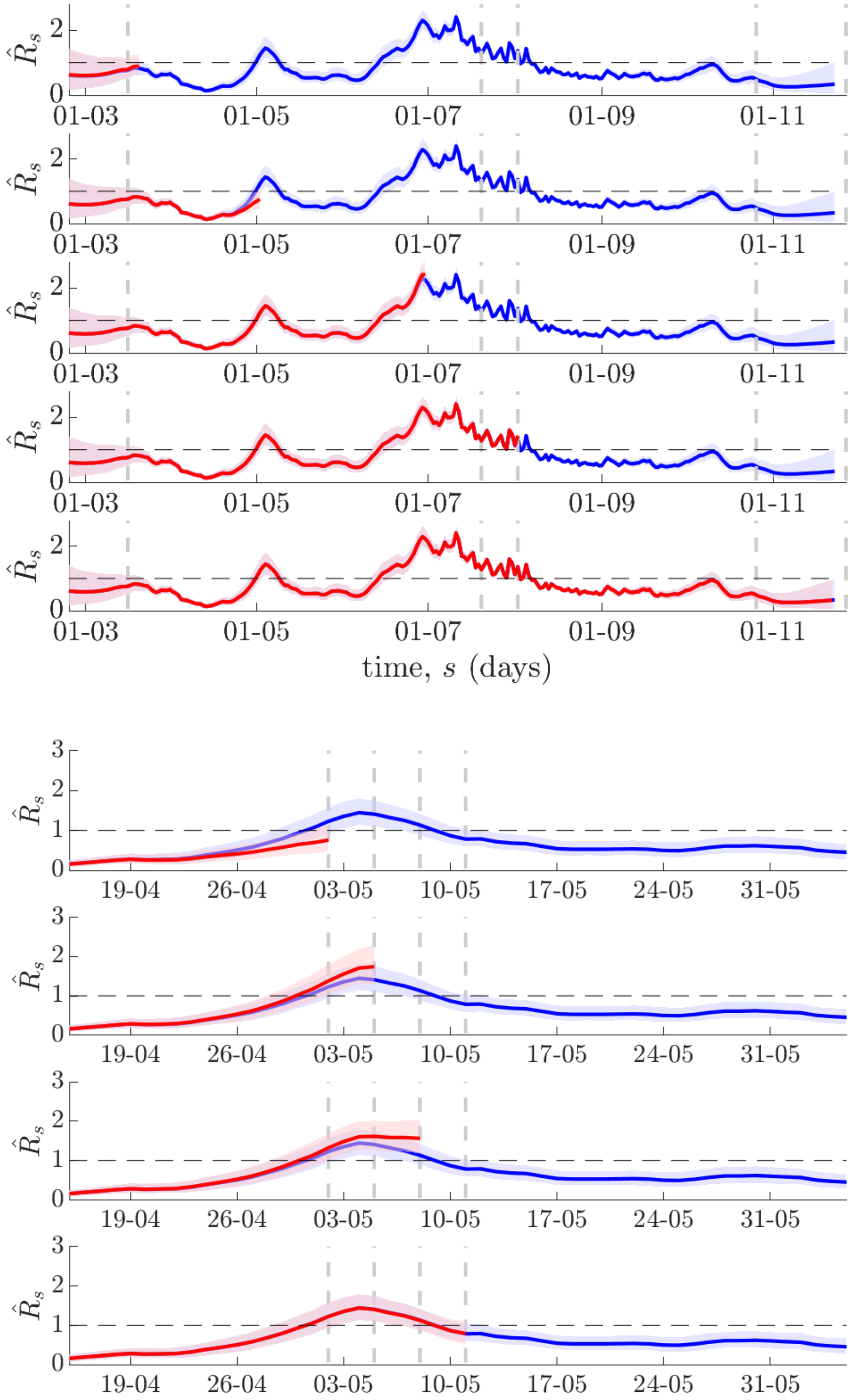

**Figure G: Real time *R*-estimates for Victoria state**. (Top) Grey lines indicate the times of key policy actions that correspond to Figure 3 of the main text. Each panel examines the smoothed, local *R*-estimates (see Methods of the main text) computed from data up to each policy action time (red with 95% confidence bands) as compared to those from Figure 3 (blue with 95% confidence bands), which processes the entire incidence curve. There is a good correspondence between the real time and retrospective estimates indicating the ability of the framework to provide early-warning signals. (Bottom) The main discrepancies are highlighted. Grey lines indicate successive 3-day releases of data. Most discrepancies (and particularly signals of resurgence) are largely resolved by the first 3-day release of data.

**Victoria: smoothed vs standard (local) *R*-estimates**

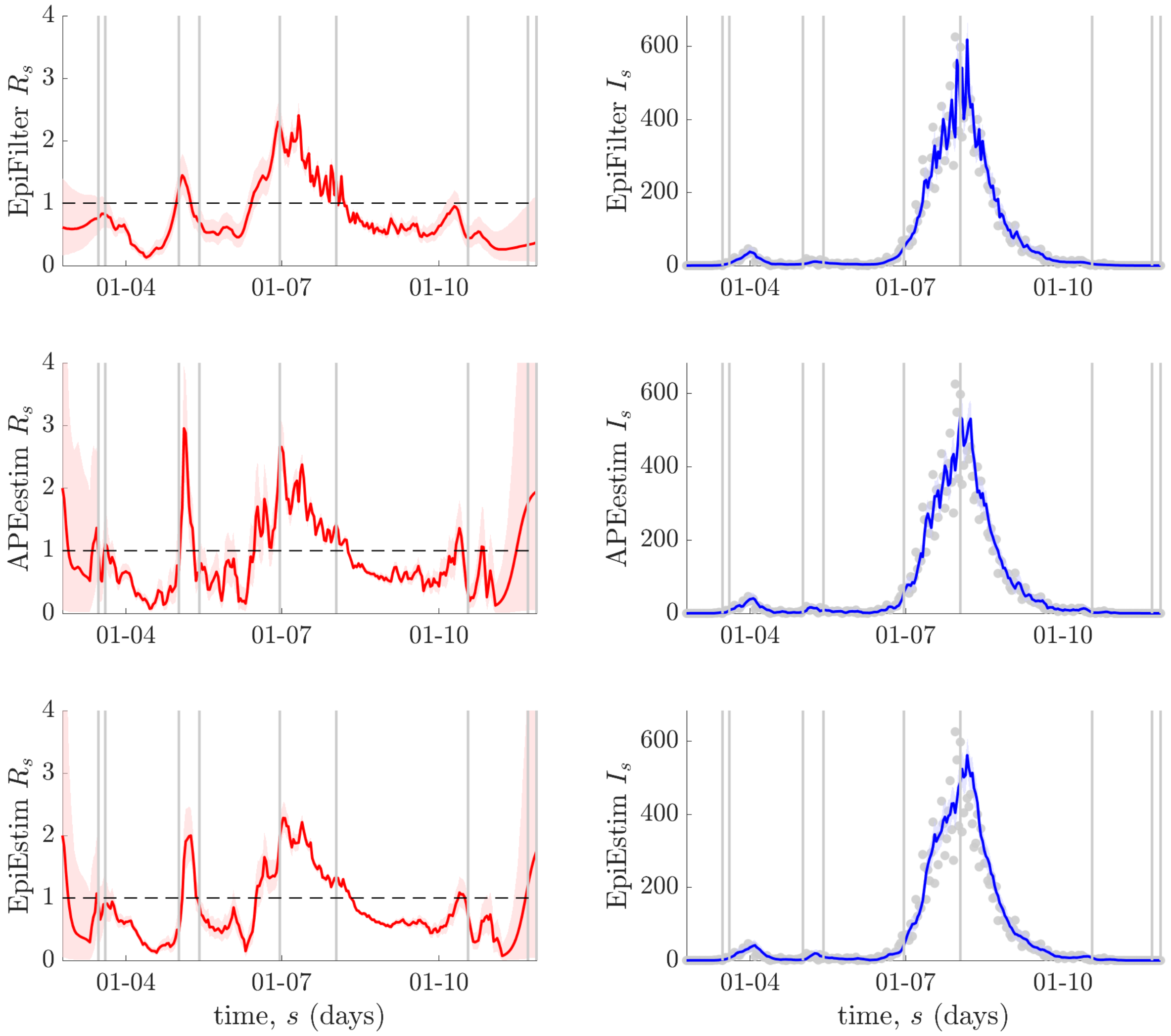

**Figure H: Local *R*-estimates for Victoria state, Australia**. Grey lines indicate the times of key policy actions as shown in Figure 3 of the main text. Left panels illustrate different *R*-estimates (red with 95% confidence bands) using *EpiFilter* (top panels, see Methods of main text), *APEestim* (middle panels, optimises window length for prediction) and *EpiEstim* (bottom panels, uses a weekly window length). Right panels provide one-step-ahead predictive fits (blue with 95% confidence bands) of the actual incidence data (grey circles) that result from the *R*-estimates of the corresponding left panel. Both *APEestim* and *EpiEstim* only use backward information, while *EpiFilter* exploits both backward and forward information, resulting in better stability and reliability, especially when incidence is small.

#### Victoria: local vs naïve (smoothed) *R*-estimates

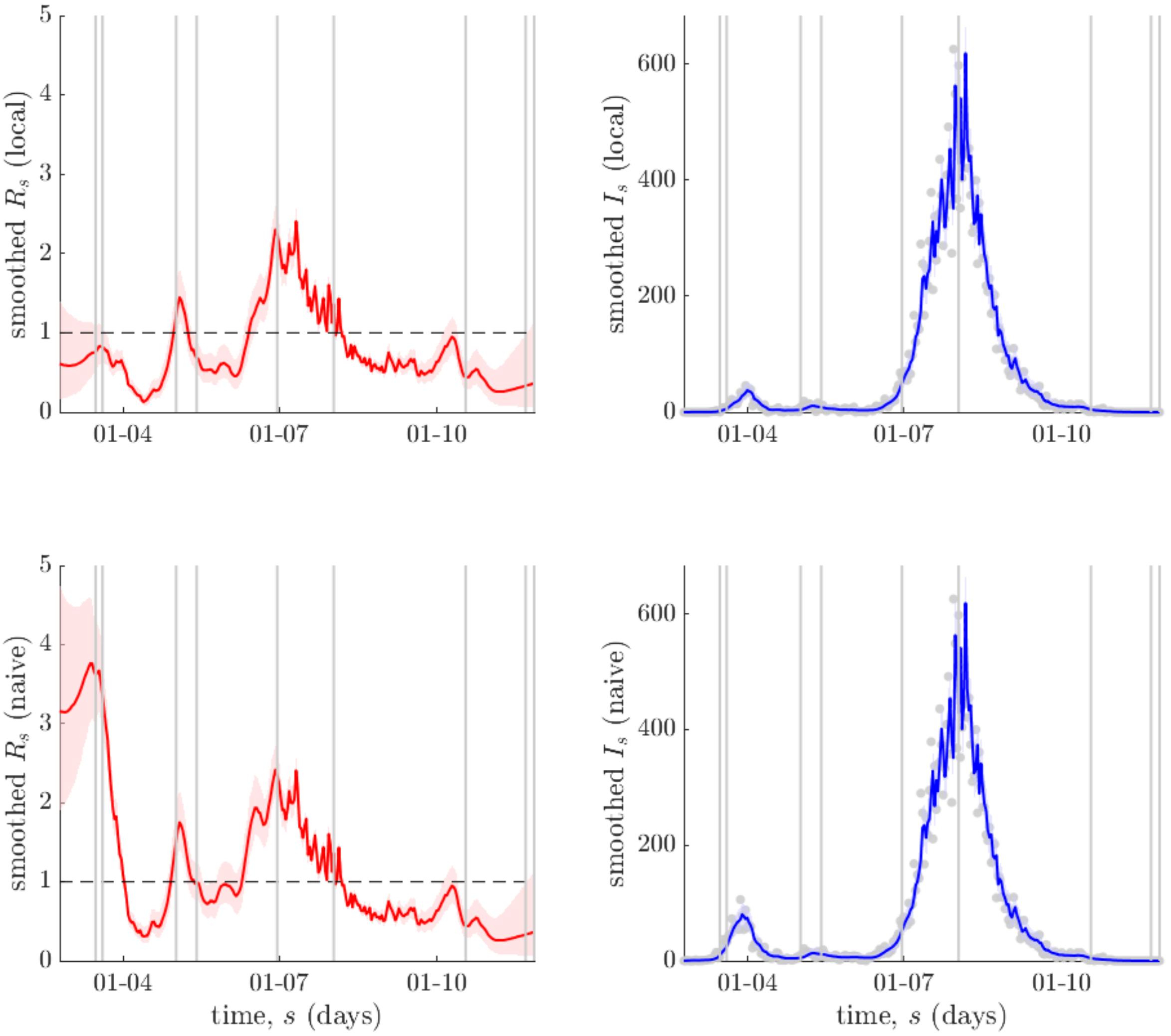

**Figure I: Smoothed *R*-estimates for Victoria state, Australia**. Grey lines indicate the times of key policy actions, corresponding to Figure 3 of the main text. Left panels illustrate the difference in smoothed *R* estimates (red with 95% confidence bands) (i.e., they use both forward and backward incidence data – see Methods of the main text and the *EpiFilter* package) using renewal models that account for (local, top panels) or ignore (naïve, bottom panels) the differing impact of local and imported cases. Naïve estimates significantly over-estimate *R*. Right panels provide one-step-ahead predictive fits of the incidence (blue with 95% confidence bands) that result from these *R*-estimates. The actual incidence data are grey circles. The fits suggest that the data are well represented by the model.

#### Victoria: local *R-Z* estimates redone with weekly averaging filter

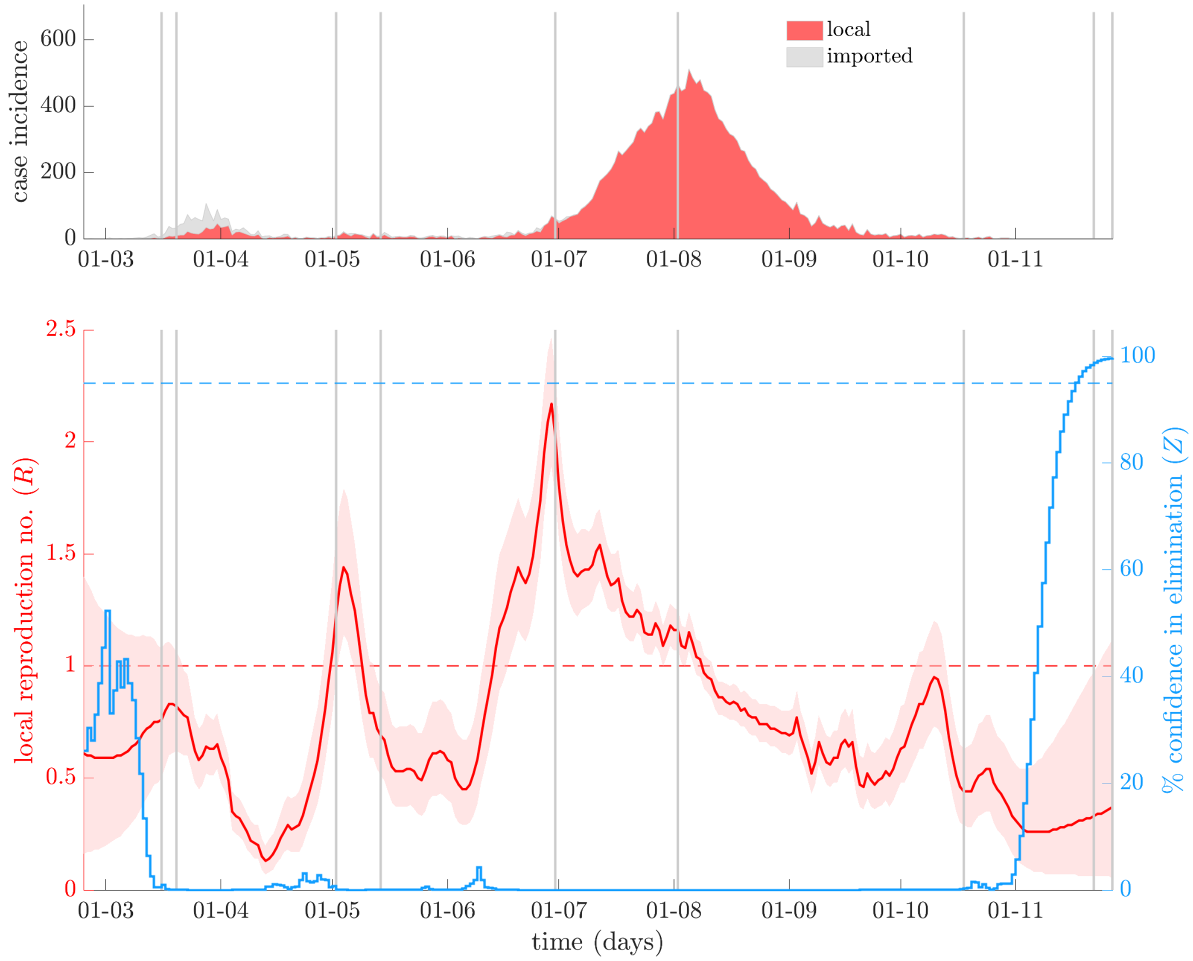

**Figure J: Local transmission dynamics of COVID-19 in Victoria state, Australia with correction for weekend reporting biases.** The top panel illustrates local (red) and imported (grey, stacked) cases by diagnosis date with a 7-day moving average filter applied throughout July to September to remove weekend variations in reporting. Vertical lines highlight important policy change-times and responses. The bottom panel presents smoothed local R-estimates (red with 95% confidence bands) and resulting Z numbers (blue) measuring the % probability of elimination using the corrected data from the top panel. This redoes the analysis presented in Figure 3 of the main text.
